# A screening tool enhances home-based identification of adolescents (aged 10-14) living with HIV in Zambia and South Africa: HPTN 071 (PopART) Study

**DOI:** 10.1101/2022.03.24.22272873

**Authors:** Mwate Joseph Chaila, David Macleod, Sten H Vermund, Moomba Mbolongwe Thornicroft, Madalitso Mbewe, Constance Mubekapi-Musadaidzwa, Abigail Harper, Albertus Schaap, Sian Floyd, Graeme Hoddinott, Richard Hayes, Sarah Fidler, Helen Ayles, Kwame Shanaube, the HPTN 071 (PopART) for Youth (P-ART-Y) Study Team

## Abstract

**Introduction:** The HPTN071 (PopART) for Youth (P-ART-Y) study evaluated the acceptability and uptake of a community-level combination HIV prevention package including universal testing and treatment (UTT) among young people in Zambia and South Africa (SA). We determined whether a four-question primary care level screening tool, validated for use in clinical settings, could enhance community (door-to-door) identification of undiagnosed HIV-positive younger adolescents (aged 10-14) who are frequently left out of HIV interventions.

**Method:** Community HIV-care Providers (CHiPs) contacted and consented adolescents in their homes and offered them participation in the PopART intervention. CHiPs used a four question-screening tool, which included: history of hospital admission; recurring skin problems; poor health in last 3 months; and death of at least one parent. A “yes” response to one or more questions was classified as being “at risk” of being HIV-positive. The data were captured through an electronic data capturing device from August 2016 to December 2017. Proportions of adolescents who were deemed “at risk” were calculated and the association of screening “at risk” with age, sex and community was tested using a chi-squared test. The adjusted odds ratio (OR) comparing the odds of testing HIV-positive if “at risk” with the odds of testing positive if “not at risk” was estimated using logistic regression.

**Results:** In our 14 study sites, 33,283 adolescents aged 10-14 in Zambia and 8,610 in SA participated in the study. About 1.3% (427/33,710) and 1.2% (106/8,610) self-reported to be HIV positive. Excluding the self-reported HIV-positive, we classified 11.3% (3,746/33,283) of adolescents in Zambia and 17.5% (1,491/8,504) in SA as “at risk”. In Zambia and SA, the “at risk” adolescents were 4.6 and nearly 16.7 times more likely to test HIV-positive compared to the “not at risk”, respectively (both p<0.001). Using the screening tool, one-third of HIV-positive adolescents could be diagnosed using just a tenth of the number of HIV tests compared to universal testing.

**Conclusion:** The screening tool may be of some value where UTT is not possible and limited resources must be prioritised toward adolescents who are more likely to be living with HIV. Further, the tool is of greater value in settings where there are more adolescents living with HIV who are undiagnosed. However, given our goal is to identify and treat all ALHIV, as well as link all HIV uninfected young people to prevention services, this screening tool should not be a substitute for UTT in community settings.

**Clinical Trial Number:** **NCT01900977**

## INTRODUCTION

HIV infection among adolescents remains a challenge, with ≈1.8 million adolescents estimated to be living with HIV globally in 2015 (1). In Africa, HIV/AIDS is now the leading cause of death among adolescents, notably among girls, and is ranked second globally next to unintentional injuries (1,2). Furthermore, an estimated 250,000 adolescents aged 15-19 become newly HIV-positive annually (3). Compared to adults, older adolescents, and children/infants, little is known about the burden of HIV and AIDS among young adolescents (aged 10-14 years) as data on them are rarely collected, and the statistics that are reported aggregate them with older youth. The other challenge is that, in many high burden countries, this age group requires parental consent for the provision of sexual and reproductive health service(1,4,5).

Another reason that very few studies report HIV testing uptake, knowledge levels, and behaviours in 10-14 year olds is that this age group is rarely a target group for HIV testing (6,7). This has likely disadvantaged programmatic planning for HIV testing, treatment and prevention services for young adolescents(7). Although estimates of the HIV prevalence in this 10–14-year-old age group was limited, among 15–19-year-olds in Zambia, in 2018, the prevalence was estimated to be 2.6% and 1.2% among females and males respectively (8). In South Africa in the same age group the estimates were 5.9% in females and 4.1% in males in 2016 (9).

Older children and adolescents infected perinatally often remain undiagnosed with HIV-infection and as such would not receive antiretroviral therapy (ART)(10–13). Efforts to identify such undiagnosed adolescents have been implemented in sub-Saharan Africa; a systematic review and other studies have shown that while provider-initiated testing and counselling (PITC) will capture some youth, additional interventions at the community level are needed to reach more youth, for example, through family-based testing, index testing with parents or family members as entry points, and distribution of HIV self-testing kits (12,14,15).

In 2011, Ferrand et al. developed a primary care level algorithm (screening tool) for identifying adolescents living with HIV in populations at high risk of vertical transmission. The screening tool consisted of five basic questions to identify adolescents aged 10-19 attending primary care facilities, who were more likely to be at risk of being HIV-positive in Zimbabwe(16). The questions were further reduced from five to four for use in children and adolescents aged 6-15 years(10). These four questions were later applied in community settings of Zimbabwe for children and adolescents aged 8-17 years (17).

Using the data collected during the PopART for Youth (P-ART-Y) study, we sought to determine whether this four-question screening tool could efficiently identify undiagnosed adolescents living with HIV (ALHIV) aged 10-14 in community-level interventions in Zambia and South Africa. The design of the PopART intervention, which was conducted through a door-to-door approach, allowed us to address this question using a very large sample of adolescents.

## METHODS

### Trial design and setting

The PopART for Youth (P-ART-Y) study aimed to determine the acceptability and uptake of HIV testing among adolescents and young adults aged 10-24 years (young people) and was implemented from October 2015 to December 2017, the main findings from the study have been published elsewhere (13,18,19). The P-ART-Y study was nested within the HPTN071 (PopART) trial, a three-arm cluster-randomized trial conducted in 12 communities in Zambia and nine communities in South Africa which was implemented from November 2013 to December 2017 (13). The HPTN071 (PopART) trial was aimed at assessing the impact of a combination HIV prevention package, including universal HIV testing and treatment (UTT), on community-level HIV incidence(20). The 21 communities were divided into seven triplets based on geography and baseline HIV prevalence. Each community in the triplet was randomly assigned to one of the three arms. Arm A received the full PopART HIV intervention which included home-based HIV testing services (HTS), immediate access to antiretroviral therapy (ART) regardless of CD4+ cell count for all diagnosed with HIV and offer of HIV prevention services like distribution of condoms, lubricants and referral for voluntary medical male circumcision (VMMC) in HIV negative men. Arm B received the full PopART intervention, but ART was initially provided according to national ART guidelines. Arm C was the standard-of-care arm in which the national ART guidelines were followed without PopART intervention enhancements. Home-based HTS was conducted by trained community health workers called Community HIV-care Providers (CHiPs) through a finger-prick rapid test using Alere Determine™ HIV-1/2 as the screening test and Uni-Gold™ Recombigen© HIV-1/2 as the confirmatory test. Further details of the main trial are described elsewhere (20). Arms B and C moved to a universal treatment policy as of May 2016 in Zambia and November 2016 in South Africa following World Health Organization (WHO) recommendations (21).

The P-ART-Y study was implemented in three phases: the qualitative baseline study and collection of data from the on-going HPTN071 (PopART) trial (phase 1) (20), addition of youth-targeted interventions in communities (phase 2) which included integration of school based intervention in all study communities; and a cross-sectional survey to measure the knowledge of HIV status in Arm C so that it could be compared with Arms A and B to see how the intervention changed knowledge of HIV status (phase 3) (13).

In Zambia, the P-ART-Y study was implemented across 4 provinces (Central, Copperbelt, Lusaka and Southern) and 6 districts (Choma, Kabwe, Kitwe, Livingstone, Lusaka, and Ndola). In South Africa, the study was conducted in the Western Cape Province (Cape Metro and Cape Winelands districts). Communities were defined as the catchment population of a local health facility (through which ART was delivered), including all schools in the selected area (13).

#### The PopART intervention

The HPTN071 (PopART) combination HIV prevention package was delivered by CHiPs via a door-to-door approach, with treatment and care-related services provided by local government clinics (22). The intervention was offered in all intervention communities (both Arm A and Arm B), eight in Zambia and six in South Africa, via three data collection rounds (see section on data collection rounds). Each data collection round lasted approximately 15 to 20 months. The P-ART-Y study was implemented in the second and third data collection rounds of the HPTN071 (PopART) study which ranged from October 2015 to December 2017.

The CHiPs teams enumerated all household members in the community including those who were reported absent. They offered home-based HIV testing services (HTS), support for linkage-to-care of all individuals diagnosed HIV-positive, ART adherence support, referral of HIV-negative males for voluntary medical male circumcision (VMMC), screening for tuberculosis (TB), and screening for sexually transmitted infections (STI). CHiPs worked in pairs within an allocated zone (450–500 households) of a given community. The CHiPs offered HIV testing to eligible participants, namely those individuals who accepted the PopART intervention and did not self-report as HIV-positive. Throughout each annual round, CHiPs arranged repeat household visits to monitor linkage to services, offering HTS for those absent at previous visits. The CHiPs recorded basic data on the household and all household members on an electronic data capture device, as well as more detailed data such as HIV test history and HIV test results of all consenting participants. Details of the PopART intervention, informed consent, and HTS are described elsewhere (13,20,22,23).

#### The P-ART-Y Screening tool

We asked four questions to parents, guardians, or other caretakers of adolescents aged 10-14 years. The questions were drawn from a previously developed primary care-level screening algorithm(10), and were asked prior to the offer of an HIV test. A “yes” response to any one or more of the following questions meant that adolescent was considered to have an increased risk of being HIV-positive (“at risk”):

1. Has the child ever been admitted to hospital?
2. Does the child have recurring skin problems?
3. Are one or both parents of the child deceased?
4. Has the child had poor health in the past 3 months?

We did not place an emphasis on following up adolescents who were classified as “not at risk” and were not at home at the time of the CHiPs’ visit. However, for those that were classified as being “at risk”, an average of three attempts were made to find and test them.

The objective of this study was to assess the effectiveness of using this screening tool to identify 10–14-year-olds at greater risk of being HIV-positive. The adolescent’s parents or guardian needed to provide written consent to allow enrolment into the PopART intervention. All children aged 10-14 years old who participated in the intervention, did not report to CHiPs being HIV-positive and had a result from a CHiP administered rapid HIV test were included in the analysis. This information was only collected in the PopART intervention arms (A and B), no data from arm C is included.

#### Data Collection Rounds

The data collection rounds were periods in which the CHiPs in each community collected data. These happened in three overlapping periods. The three rounds were: Round 1 (R1): November 2013 –June 2015; Round 2 (R2): June 2015 – October 2016; and Round 3 (R3): August 2016 – December 2017. We used data from the period August 2016 to December 2017 (R3) as the data were the most complete in both countries for that round.

#### Study objective, participant eligibility, data collection and analysis

The CHiPs recorded all household data in an electronic data capture device. Proportions of adolescents who were deemed “at risk” were calculated and the association of screening “at risk” with age, sex and community was tested using a chi-squared test. The adjusted odds ratio (OR) comparing the odds of testing HIV-positive if “at risk” with the odds of testing positive if “not at risk” was estimated using a logistic regression. This regression included a random effect for CHiPs work zones, and the model was also adjusted for the participants’ age, sex and community.

To assess the sensitivity, specificity, and the positive and negative predictive values of the screening tool, both the screening result and the HIV test result (the gold standard) needed to be available, and this was only available for those who accepted HIV testing. So, the sample used for this analysis was among those who accepted an HIV test from the CHiPs. To allow for the differences in uptake of HIV tests between those “at risk” and “not at risk”, the subsequent analysis was weighted using inverse-probability weighting to take into account the fact that we had the true HIV status of more individuals in the “at risk” group. The probability of testing was estimated using logistic regression with “accepted test” as the outcome and “at risk” as the exposure; then the inverse of this probability was applied when attempting to estimate sensitivity and specificity. It was assumed that acceptance of testing was independent of HIV status among those in the same risk group, i.e., within each risk group the prevalence of HIV was the same among those who accepted testing and those who declined. The prevalence of HIV-positive status among those who were not previously known to be HIV-positive was estimated in each country, using the inverse probability weights to account for the difference in uptake of testing by risk group. We further extrapolated the results to a hypothetical population of 10,000 adolescents aged 10-14 to estimate the number of HIV-positive adolescents identified by using the screening tool. Stata version 16 was used for all data management and analysis.

#### Ethical approval

Ethics approval was obtained from the ethics committees of the University of Zambia, Stellenbosch University and the London School of Hygiene and Tropical Medicine. Permission to conduct the study was received from the Zambian Ministry of Health and the Western Cape Department of Health in South Africa. We sought informed assent and consent from adolescents and their guardians respectively (13).

## RESULTS

### Participation

There were 49,048 adolescents aged 10-14 enumerated within the eight Zambian communities during R3, of whom 33,710 (68.7%) participated in the PopART intervention. Absence from the household at the time of the CHiPs team visit was the primary reason for non-participation (90.5% of all non-participants). Among these adolescents, 427 (1.3%) self-reported being HIV-positive leaving 33,283 participants eligible for HIV testing by CHiPs (Figure 1a).

**Figure 1a:**
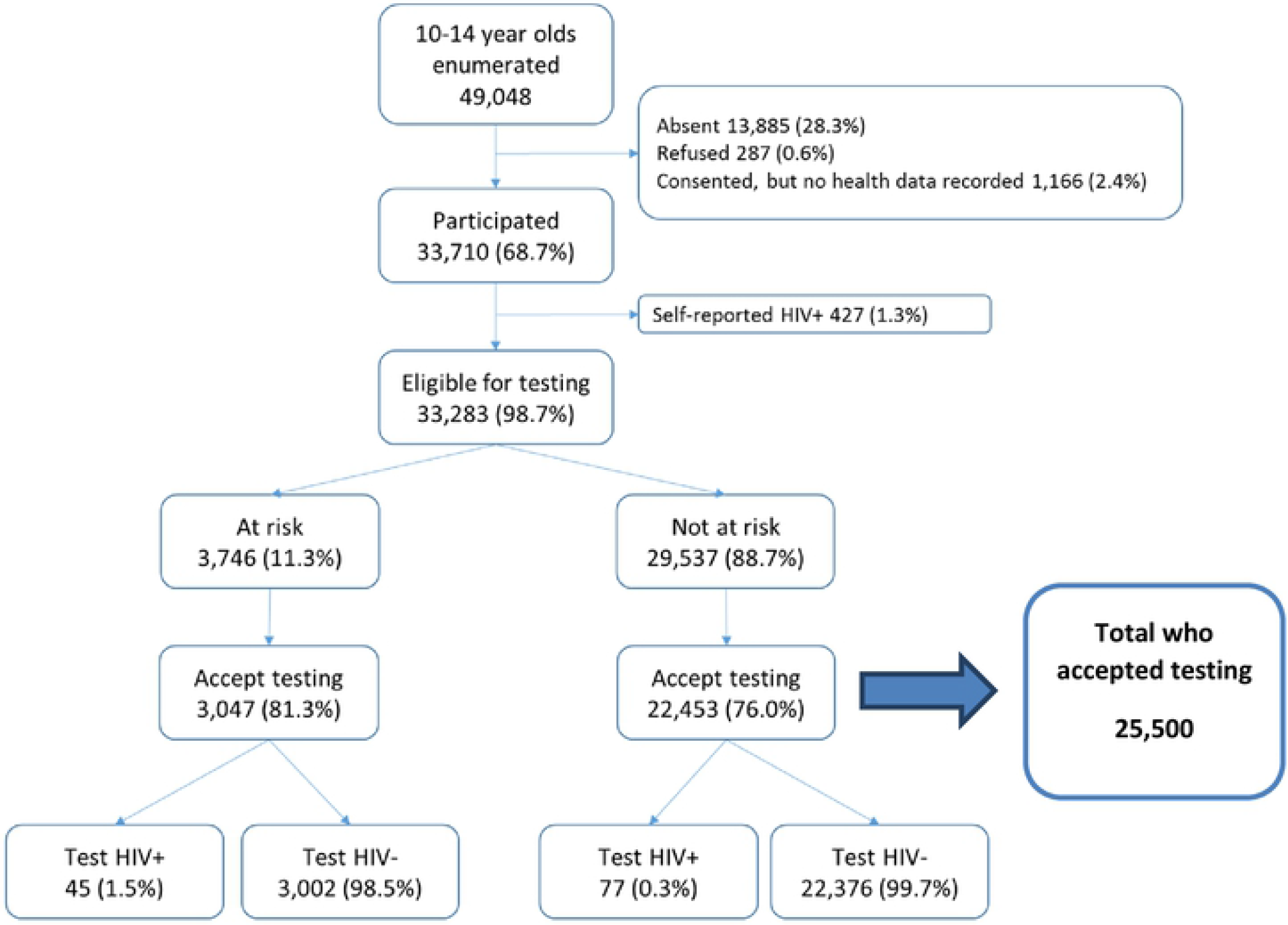
Flow chart for the HIV testing cascade in adolescents aged 10-14 years (Zambia) in the P-ART-Y sub-study of HPTN 071 (PopART)

In South Africa 16,956 adolescents were enumerated with 8,610 (50.8%) participating in the intervention. Again, most non-participation (84.8%) was due to absence from the household. There were 106 (1.2%) participants who self-reported living with HIV, resulting in 8,504 participants eligible for HIV testing in South Africa (Figure1b).

**Figure 1b:**
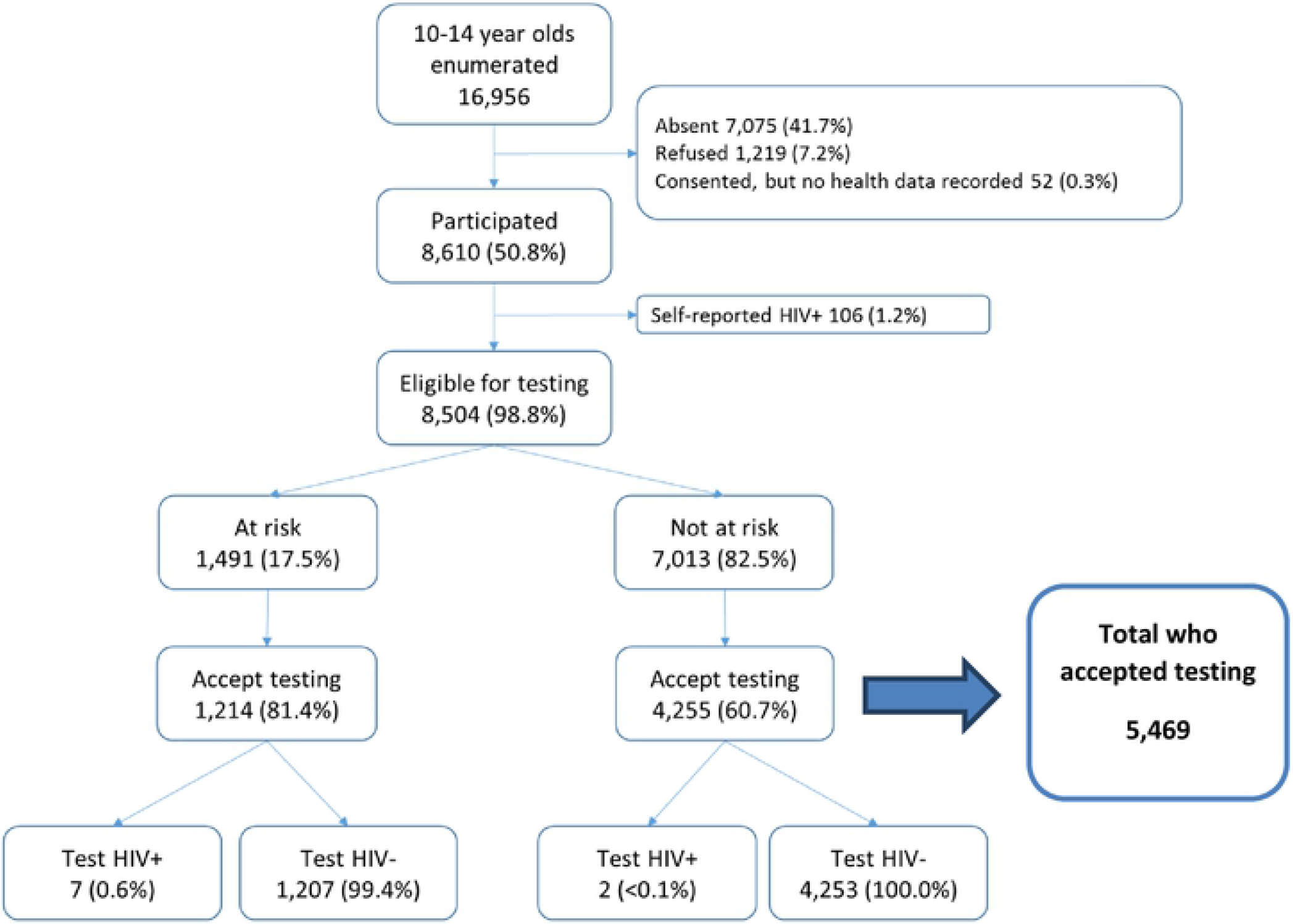
Flow chart for the HIV testing cascade in adolescents aged 10-14 years (South Africa) in the P-ART-Y sub-study of HPTN 071 (PopART)

### Screening questions and uptake of HIV testing

In R3, Zambia had 11.3% (3,746/33,283) of eligible participants classified as being “at risk.” Acceptance of HIV testing was 81.3% (3,047/3,746) in the “at risk” and 76.0% (22,453/29,537) in the “not at risk” groups (figure 1a). In South Africa, 17.5% (1,491/8,504) were classified as “at risk”. Acceptance of HIV testing was 81.4% (1,214/1,491) in the “at risk” and 60.7% (4,255/7,013) in the “not at risk” groups (Figure 1b).

In both countries, there was no substantial difference between the proportion of adolescent boys and girls who were classified “at risk”. In Zambia there was a small increase in the proportion of those who were “at risk” with increase in age; however, the pattern was not the same in South Africa, where adolescents aged 10 had the highest proportion of being “at risk” and those aged 11-14 had a lower proportion of being at “at risk”. In both countries, there were marked differences between communities (Tables 1a and 1b) which is expected in community-randomized trials where communities are known to be different (24).

**Table 1a:** Demographics among all 10-14 year olds screened who did not self-report being HIV-positive (Zambia) in the P-ART-Y sub-study of HPTN 071 (PopART)

|  |  | N (%) of all 10-14 year olds eligible for HIV testing within each category* |  | N (%) "at risk" among those eligible for testing within each category |  | p-value† |
| --- | --- | --- | --- | --- | --- | --- |
| **Overall** |  |  |  |  |  |  |
|  | All | 33,283 | 100.0% | 3,746 | 11.3% |  |
| **Sex** |  |  |  |  |  |  |
|  | M | 15,206 | 45.7% | 1,716 | 11.3% | 0.874 |
|  | F | 18,077 | 54.3% | 2,030 | 11.2% |  |
| **Age** |  |  |  |  |  |  |
|  | 10 | 7,525 | 22.6% | 729 | 9.7% | <0.001 |
|  | 11 | 6,631 | 19.9% | 723 | 10.9% |  |
|  | 12 | 6,673 | 20.0% | 729 | 10.9% |  |
|  | 13 | 6,396 | 19.2% | 794 | 12.4% |  |
|  | 14 | 6,058 | 18.2% | 771 | 12.7% |  |
| **Community** |  |  |  |  |  |  |
|  | 1 | 3,572 | 10.7% | 466 | 13.0% | <0.001 |
|  | 2 | 1,400 | 4.2% | 106 | 7.6% |  |
|  | 5 | 5,693 | 17.1% | 849 | 14.9% |  |
|  | 6 | 3,267 | 9.8% | 262 | 8.0% |  |
|  | 8 | 6,577 | 19.8% | 606 | 9.2% |  |
|  | 9 | 7,715 | 23.2% | 1034 | 13.4% |  |
|  | 10 | 2,674 | 8.0% | 223 | 8.3% |  |
|  | 11 | 2,385 | 7.2% | 200 | 8.4% |  |

† p-value from chi-squared test

**Table 1b:** Demographics among all 10-14 year olds screened, who did not self-report being HIV-positive (South Africa) in the P-ART-Y sub-study of HPTN 071 (PopART)

|  |  | N (%) of all 10-14 year olds<br>eligible for HIV testing within<br>each category* |  | N (%) "at risk" among those<br>eligible for testing within each<br>category |  | p-value† |
| --- | --- | --- | --- | --- | --- | --- |
| **Overall** |  |  |  |  |  |  |
|  | All | 8,504 | 100.0% | 1,491 | 17.5% |  |
| **Sex** |  |  |  |  |  |  |
|  | M | 3,954 | 46.5% | 689 | 17.4% | 0.808 |
|  | F | 4,550 | 53.5% | 802 | 17.6% |  |
| **Age** |  |  |  |  |  |  |
|  | 10 | 1,871 | 22.0% | 365 | 19.5% | 0.144 |
|  | 11 | 1,754 | 20.6% | 291 | 16.6% |  |
|  | 12 | 1,680 | 19.8% | 284 | 16.9% |  |
|  | 13 | 1,635 | 19.2% | 284 | 17.4% |  |
|  | 14 | 1,564 | 18.4% | 267 | 17.1% |  |
| **Community** |  |  |  |  |  |  |
|  | 13 | 1,577 | 18.5% | 319 | 20.2% | <0.001 |
|  | 14 | 524 | 6.2% | 59 | 11.3% |  |
|  | 16 | 2,707 | 31.8% | 484 | 17.9% |  |
|  | 18 | 1,501 | 17.7% | 394 | 26.2% |  |
|  | 19 | 917 | 10.8% | 137 | 14.9% |  |
|  | 20 | 1,278 | 15.0% | 98 | 7.7% |  |

† p-value from chi-squared test

### HIV test results by risk group

In Zambia 122/25,500 (0.5%) of adolescents who tested with CHiPs had a positive result, and in SA it was 9/5,469 (0.2%). Both Zambia and South Africa had a higher proportion with a positive test result in the “at risk” group compared to the “not at risk.” In Zambia, it was 1.5% (45/3,047) in the “at risk” and 0.3% (77/22,453) among those in the “not at risk” group. In South Africa with fewer newly-diagnosed individuals found, the proportion testing positive was 0.6% (7/1,214) in the “at risk” group and 0.0005% (2/4,253) in the “not at risk” group.

In both countries, there was strong evidence of an association between screening “at risk” and testing positive for HIV. In Zambia those who screened “at risk” were estimated to have 4.6 times the odds of testing HIV-positive compared to those “not at risk”, after adjusting for age, sex and community (95%CI: 3.1-6.6, p<0.001). In South Africa the estimated increase in odds was 16.7 times among those “at risk” (95% CI: 3.4-83.3, p<0.001) (Table 2).

**Table 2:**
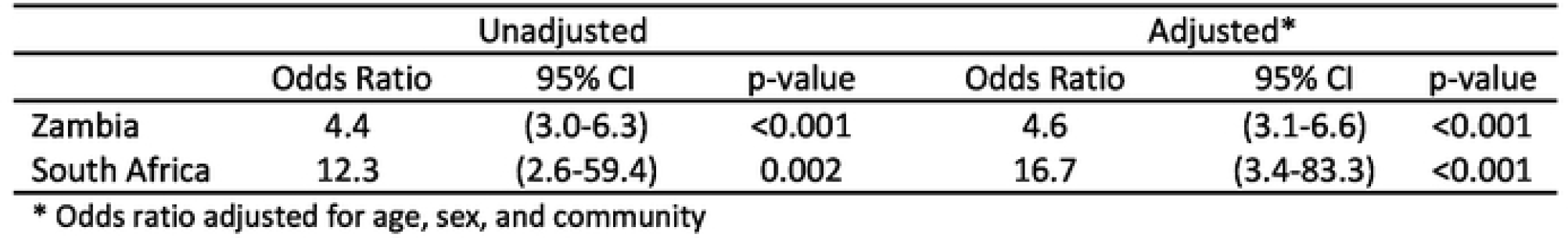
Association between screening “at risk” and testing HIV positive In the P-ART-Y sub-study of HPTN 071 (PopART)

|  | Unadjusted |  |  | Adjusted* |  |  |
| --- | --- | --- | --- | --- | --- | --- |
|  | Odds Ratio | 95% CI | p-value | Odds Ratio | 95% CI | p-value |
| Zambia | 4.4 | (3.0-6.3) | <0.001 | 4.6 | (3.1-6.6) | <0.001 |
| South Africa | 12.3 | (2.6-59.4) | 0.002 | 16.7 | (3.4-83.3) | <0.001 |

\* Odds ratio adjusted for age, sex, and community

### Sensitivity and Predictive Value of the Screening tool

The sensitivity, specificity and the positive and negative predictive values (25) were estimated among those who accepted an HIV test from the CHiPs (25,500 in Zambia; and 5,469 in South Africa) (Tables 3a and 3b).

**Table 3a:** Sensitivity and specificity, positive predictive value (PPV), negative predictive value (NVP), number needed to test (NNT) to identify 1 HIV-infected after application of screening tool (Zambia) in the P-ART-Y sub-study of HPTN 071 (PopART)

|  | Sensitivity (%) | Specificity (%) | PPV (%) | NPV (%) | NNT |
| --- | --- | --- | --- | --- | --- |
| **Screening Method** |  |  |  |  |  |
| Universal testing | 100 | 0 | 0.5 (0.4, 0.6) | N/A | 213 (178, 254) |
| Screened "at risk" | 35.3 (27.3, 44.2) | 88.9 (88.5, 89.2) | 1.5 (1.1, 2.0) | 99.7 (99.6, 99.7) | 68 (51, 91) |
| Alternative screening | 33.8 (25.9, 42.6) | 91.4 (91.0, 91.7) | 1.8 (1.3, 2.4) | 99.7 (99.6, 99.7) | 55 (41, 74) |
| **Individual Questions** |  |  |  |  |  |
| Admitted to hospital | 5.5 (2.6, 11.2) | 96.6 (96.4, 96.9) | 0.8 (0.4, 1.6) | 99.5 (99.4, 99.6) | 130 (62, 273) |
| Skin problems | 12.6 (7.8, 19.7) | 99.1 (99.0, 99.2) | 6.1 (3.8, 9.8) | 99.6 (99.5, 99.7) | 16 (10, 27) |
| Parent died | 24.3 (17.5, 32.7) | 92.8 (92.5, 93.1) | 1.6 (1.1, 2.2) | 99.6 (99.5, 99.7) | 64 (45, 90) |
| Poor health | 12.6 (7.8, 19.7) | 99.0 (98.8, 99.1) | 5.5 (3.4, 8.8) | 99.6 (99.5, 99.7) | 18 (11, 30) |

**Table 3b:** Sensitivity and specificity, positive predictive value (PPV), negative predictive value (NVP), number needed to test (NNT) to identify 1 HIV-infected after application of screening tool (South Africa) in the P-ART-Y sub-study of HPTN 071 (PopART)

|  | Sensitivity (%) | Specificity (%) | PPV (%) | NPV (%) | NNT |
| --- | --- | --- | --- | --- | --- |
| **Screening Method** |  |  |  |  |  |
| Universal testing | 100 | 0 | 0.1 (0.1, 0.3) | N/A | 715 (370, 1382) |
| Screened "at risk" | 72.3 (26.8, 94.9) | 82.5 (81.6, 83.4) | 0.6 (0.3, 1.2) | 100.0 (99.8, 100.0) | 173 (83, 364) |
| Alternative screening | 51.6 (17.0, 84.8) | 86.5 (85.6, 87.3) | 0.5 (0.2, 1.3) | 99.9 (99.8, 100.0) | 188 (79, 452) |
| **Individual Questions** |  |  |  |  |  |
| Admitted to hospital | 20.7 (3.5, 65.0) | 93.5 (92.9, 94.1) | 0.4 (0.1, 1.8) | 99.9 (99.7, 99.9) | 224 (56, 900) |
| Skin problems | 31.0 (7.3, 71.8) | 93.9 (93.3, 94.5) | 0.7 (0.2, 2.2) | 99.9 (99.8, 100.0) | 141 (46, 439) |
| Parent died | 20.7 (3.5, 65.0) | 93.3 (92.7, 93.9) | 0.4 (0.1, 1.7) | 99.9 (99.7, 99.9) | 232 (58, 930) |
| Poor health* | ... | 97.5 (97.1, 97.8) | ... | 99.9 (99.7, 99.9) |  |

\*In SA, among those who tested HIV-positive none answered yes to the question "has the child had poor health in the past 3 months"

The screening tool had an estimated sensitivity of 35.3% (95% CI: 27.3%-44.2%) and an estimated specificity of 88.9% (95% CI: 88.5%-89.2%). This resulted in an estimated positive predictive value (PPV) of 1.5% (95% CI: 1.1%-2.0%), giving a number needed to test (NNT) to obtain, on average, one HIV-positive test result of 68 (95% CI: 51-91). In South Africa, the estimated sensitivity was higher at 72.3% (95% CI: 26.8%-94.9%), but this estimate was very imprecise given the very low numbers of undiagnosed HIV-positive 10–14-year-olds. The specificity was estimated to be 82.5% (95% CI: 81.6%-83.4%) and the PPV was 0.6% (95% CI: 0.3%-1.2%) resulting in an estimated NNT of 173 (95% CI: 83-364).

In Zambia, if 10,000 adolescents aged 10-14 accepted HIV testing (among participants who did not self-report as HIV-positive), we would expect to find 47 testing HIV-positive. If we first applied the screening tool to these 10,000 individuals, we would expect 1,126 of them to answer yes to ≥1 of the questions, and 17 of these to test HIV-positive. So, by screening first we would expect to find 17/47 (35%) of those who were HIV-positive but we would reduce the number of HIV tests done by 8,874(89% fewer tests). In South Africa, our best estimate is that out of 10,000 adolescents, we would expect 14 to be HIV-positive and using screening we would expect to identify 10/14 of these (72%) and reduce the number of tests carried out by 8,247 (82.5%); however, the estimate of sensitivity is imprecise so the inference that can be drawn from this is limited.

The estimated proportion who were HIV-positive among those who did not self-report HIV-positive was 0.5% (95% CI: 0.4%-0.6%) in Zambia and 0.1% (95% CI: 0.1%-0.3%) in South Africa. These values are interpreted as PPV of universal testing, i.e., classifying every 10-14 year-old who did not self-report being HIV living with HIV as “at risk” and offering tests to all. This gives an NNT for universal HIV testing in Zambia of 213 (95% CI: 178-254) and 715 (95% CI: 370-1,382) in South Africa.

### Screening questions

The individual questions were investigated to identify if all were appropriate screening questions in this setting. In Zambia, ≈1% answered yes to whether they had recurring skin problems or poor health in the last three months. About 3.5% had ever been admitted to hospital and ≈7% had a parent who had died. In South Africa the prevalence of ever being admitted to hospital (6%) and having skin problems (6%) was higher than in Zambia. The sensitivity and specificity for each of these questions were separately estimated (Table 3). In Zambia, the question “has your child ever been admitted to hospital?” had a low sensitivity, so the sensitivity and specificity of a hypothetical alternative-screening tool was estimated (alternative screening row on table 3a). It was found that by excluding this question in Zambia would only lead to a small (2-4%) reduction in the sensitivity of the screening, for a similarly small reduction in the estimated NNT which would range from 4-13.

## DISCUSSION

This study explored the use of a validated four-question screening tool to identify HIV-positive adolescents aged 10-14 in a community setting in Zambia and South Africa. The study took advantage of the large community intervention that was implemented in the HPTN071 (PopART) trial. A key finding in both countries was that those who screened “at risk” were more likely to test HIV-positive compared to those who were “not at risk”. However, the sensitivity of the screening tool was only about 35% in Zambia and somewhat higher in South Africa at 72%. These findings are broadly similar to what was found in community settings in Zimbabwe where the sensitivity of the screening tool was 56.3% (95% CI: 44.0–68.1%) (17), much lower than when done in clinical settings (10,16). This difference could be explained by the fact that when the screen was applied to ill children who visited the health facilities, the positive predictive value was much higher, as HIV prevalence was likely higher in this group.

This study showed that testing just over a tenth of all adolescents, would identify about a third of the HIV-positive cases if the screening tool were applied in Zambia. Of the four questions, death of a parent was the most sensitive question for testing positive for HIV, with a sensitivity of over 20% in both countries. This is similar to what was found in the clinical settings of Zimbabwe (10,16). Previous hospital admission was the least predictive element. Our results show that in a setting similar to that in R3 in Zambia, if the question on previous hospital admission was dropped when screening 10,000 adolescents aged 10-14, we would expect to miss three additional HIV-positive adolescents and perform 265 fewer tests (data not shown).

Since the screening tool was easy to use in both our study and that of Bandason et al. (10,17) it could be sustainable alternative to universal testing, if there are insufficient resources to test all 10-14 year olds. The screening tool was initially designed to identify children and adolescents infected through mother-to-child-transmission presenting at health facilities. Therefore, the PPV of this tool is likely to drop over time with the move to universal treatment for people living with HIV and improved strategies for prevention-of-mother-to-child-transmission (10,16). A tool with a higher sensitivity would be desirable for community screening (17). The values of testing HIV negative is that it’s a pathway for these adolescents in accessing HIV presentation services (26).

A systematic review highlighted that innovative population-based HTC strategies that could easily be brought to scale were needed that could be implemented effectively, efficiently and economically at a population level (11,26). Similarly WHO cited the need for research in pre-HIV testing screening questions to identify at risk populations (27). Our study found that this screening tool, when used in a community setting, had a low sensitivity, missing two-thirds of the ALHIV in our population. It did however reduce the number of tests to be performed by around 90% compared with universal testing, so there would be a reduced financial cost associated with performing this pre-screening but a great cost in terms of missed diagnoses and therefore the tool should not be a replacement for universal testing of adolescents.

The strengths of this study included the very large number of young adolescent study participants, unprecedented in prior HIV research; use of the community setting for a clinical tool; conducting the study in two countries with a high HIV prevalence; and a study population of adolescents aged 10-14, a sub-population that has never been studied at this scale. The screening tool was simple and was easily administered by community health-care workers.

The limitations are that the study was designed to focus more on finding and testing adolescents who were classified as being “at risk” compared to the “not at risk”, hence the difference in the uptake of HIV testing between these groups especially in South Africa.

## CONCLUSIONS

The screening tool may be of some value where UTT is not possible and limited resources must be prioritised toward adolescents who are more likely to be living with HIV. Further, the tool is of greater value in settings where there are more adolescents living with HIV who are undiagnosed. However, given our goal is to identify and treat all ALHIV, as well as link all HIV uninfected young people to prevention services, this screening tool should not be a substitute for UTT in community settings.

## Data Availability

The data are currently held at Zambart in Lusaka Zambia and the London School of Hygiene & Tropical Medicine in the UK and can be made available on request.

https://www.hptn.org/research/studies/hptn071

## AUTHORS’ CONTRIBUTIONS

M.J.C. took the lead on writing the paper together with K.S. and S.H.V. Statistical analysis was led by D.M and was supported by AS and SF. While MMT, MM, CMM, AH and GH contributed in the study design and implementation. RH, SFi, and HA conceived the idea of the study, provided guidance throughout the drafting phase and approved the final version of the paper.

## ACKNOWLEDGEMENTS

We would like to acknowledge the HPTN071 (PopART) and P-ART-Y study teams.

The content is solely the responsibility of the authors and does not necessarily represent the official views of the NIAID, NIMH, NIDA, PEPFAR, 3ie, the Bill & Melinda Gates Foundation or UK Aid. We are grateful to all members of the HPTN071 (PopART) and P-ART-Y Study Teams, and to the study adolescents and their communities, for their contributions to the research.

We further wish to thank Professor Tim Quinlan, a Research Associate, at Health Economics and HIV/AIDS Research Division (HEARD), University of KwaZulu-Natal, Durban in South Africa, who first supervised M.J.C during the *African Journal of AIDS Research* (AJAR) writing workshop in Cape Town, South Africa.

HPTN071 was sponsored by the National Institute of Allergy and Infectious Diseases (NIAID) under Co-operative Agreements UM1-AI068619, UM1-AI068617, and UM1-AI068613, with funding from the U.S. President’s Emergency Plan for AIDS Relief (PEPFAR). Additional funding was provided by the International Initiative for Impact Evaluation (3ie) with support from the Bill & Melinda Gates Foundation, as well as NIAID, the National Institute on Drug Abuse (NIDA) and the National Institute of Mental Health (NIMH), all part of the U.S. National Institutes of Health (NIH). RH and SFl receive funding from the UK Medical Research Council (MRC) and the UK Foreign, Commonwealth and Development Office (FCDO) under the MRC/FCDO Concordat agreement and is also part of the EDCTP2 programme supported by the European Union. Grant Ref: MR/R010161/1.

The P-ART-Y study was funded by the Evidence for HIV Prevention in Southern Africa (EHPSA), a UK aid programme managed by Mott MacDonald.

## REFERENCES

1. UNAIDS. Ending the AIDS epidemic for adolescents, with adolescents. 2016;36.

2. WHO. Adolescent and young adult health [Internet]. 2021 [cited 2021 Nov 11]. Available from: https://www.who.int/news-room/fact-sheets/detail/adolescents-health-risks-and-solutions

3. UNICEF. Adolescent HIV Prevention [Internet]. UNICEF. 2018 [cited 2018 Oct 28]. Available from: https://data.unicef.org/topic/hivaids/adolescents-young-people/

4. Idele P, Gillespie A, Porth T, Suzuki C, Mahy M, Kasedde S, et al. Epidemiology of HIV and AIDS Among Adolescents: Current Status, Inequities, and Data Gaps. JAIDS J Acquir Immune Defic Syndr [Internet]. 2014;66. Available from: https://journals.ww.com/jaids/Fulltext/2014/07011/Epidemiology_of_HIV_and_AIDS_Among_Adolescents.2.aspx

5. Joint United Nations Programme on HIV/AIDS (UNAIDS). Global AIDS Response Progress Reporting 2014: Construction of Core Indicators for Monitoring the 2011 UN Political Declaration on HIV and AIDS. Geneva, Switzerland: UNAIDS; 2014.

6. UNICEF. A Report Card of Adolescents on Zambia. 2014.

7. Carvalho MA, Nsemukila BG. Children and Women in Zambia Update of the Situation Analysis of for Malaria Indicators). 2013; Available from: https://www.unicef.org/zambia/Updated_Situation_Analysis_of_Women_and_Children_In_Zambia.pdf

8. Zambia Statistics Agency, Ministry of Health, University Teaching Hospital Virology laboratory, ICF. Zambia Demographic and Health Survey 2018. 2019;540.

9. National Department of Health (NDoH), Statistics South Africa (Stats SA), South African Medical Research Council (SAMRC) and I. South Africa Demographic and Health Survey 2016 [Internet]. Pretoria, South Africa: Pretoria, South Africa, and Rockville, Maryland, USA: NDoH, Stats SA, SAMRC, and ICF.; 2019. Available from: https://dhsprogram.com/pubs/pdf/FR337/FR337.pdf

10. Bandason T, McHugh G, Dauya E, Mungofa S, Munyati SM, Weiss HA, et al. Validation of a screening tool to identify older children living with HIV in primary care facilities in high HIV prevalence settings. AIDS [Internet]. 2016 Mar 13;30(5):779–85. Available from: http://www.ncbi.nlm.nih.gov/pmc/articles/PMC4937807/

11. Govindasamy D, Ferrand RA, Wilmore SMS, Ford N, Ahmed S, Afnan-Holmes H, et al. Uptake and yield of HIV testing and counselling among children and adolescents in sub-Saharan Africa: a systematic review. J Int AIDS Soc [Internet]. 2015 Oct 14;18(1):20182. Available from: http://www.ncbi.nlm.nih.gov/pmc/articles/PMC4607700/

12. Lightfoot M, Dunbar M, Weiser SD. Reducing undiagnosed HIV infection among adolescents in sub-Saharan Africa: Provider-initiated and opt-out testing are not enough. PLOS Med [Internet]. 2017 Jul 25;14(7):e1002361. Available from: https://doi.org/10.1371/journal.pmed.1002361

13. Shanaube K, Schaap A, Chaila MJ, Floyd S, MacKworth-Young C, Hoddinott G, et al. Community intervention improves knowledge of HIV status of adolescents in Zambia: Findings from HPTN 071-PopART for youth study. Aids. 2017;31:S221–32.

14. Simms V, Dauya E, Dakshina S, Bandason T, McHugh G, Munyati S, et al. Community burden of undiagnosed HIV infection among adolescents in Zimbabwe following primary healthcare-based provider-initiated HIV testing and counselling: A cross-sectional survey. PLoS Med. 2017;14(7):1–15.

15. Choko AT, MacPherson P, Webb EL, Willey BA, Feasy H, Sambakunsi R, et al. Uptake, Accuracy, Safety, and Linkage into Care over Two Years of Promoting Annual Self-Testing for HIV in Blantyre, Malawi: A Community-Based Prospective Study. PLoS Med. 2015;12(9):1–21.

16. Ferrand RA, Weiss HA, Nathoo K, Ndhlovu CE, Mungofa S, Munyati S, et al. A primary care level algorithm for identifying HIV-infected adolescents in populations at high risk through mother-to-child transmission. Trop Med Int Heal [Internet]. 2011 Mar;16(3):349–55. Available from: http://www.ncbi.nlm.nih.gov/pmc/articles/PMC3132444/

17. Bandason T, Dauya E, Dakshina S, McHugh G, Chonzi P, Munyati S, et al. Screening tool to identify adolescents living with HIV in a community setting in Zimbabwe: A validation study. PLoS One [Internet]. 2018;13(10):10–5. Available from: https://doi.org/10.1371/journal.pone.0204891

18. Shanaube K, Schaap A, Hoddinott G, Mubekapi-Musadaidzwa C, Floyd S, Bock P, et al. Impact of a community-wide combination HIV prevention intervention on knowledge of HIV status among adolescents. AIDS. 2021 Feb;35(2):275–85.

19. Shanaube K, Macleod D, Chaila MJ, Mackworth-Young C, Hoddinott G, Schaap A, et al. HIV Care Cascade Among Adolescents in a “Test and Treat” Community-Based Intervention: HPTN 071 (PopART) for Youth Study. J Adolesc Heal. 2020;

20. Hayes R, Ayles H, Beyers N, Sabapathy K, Floyd S, Shanaube K, et al. HPTN 071 (PopART): Rationale and design of a cluster-randomised trial of the population impact of an HIV combination prevention intervention including universal testing and treatment - a study protocol for a cluster randomised trial. Trials. 2014;15(1):1–17.

21. WHO. Treat all people living with HIV, offer antiretrovirals as additional prevention choice for people at “substantial” risk [Internet]. WHO publications. 2015 [cited 2018 Oct 3]. Available from: http://www.who.int/mediacentre/news/releases/2015/hiv-treat-all-recommendation/en/

22. Viljoen L, Mainga T, Casper R, Mubekapi-Musadaidzwa C, Wademan DT, Bond VA, et al. Community-based health workers implementing universal access to HIV testing and treatment: Lessons from South Africa and Zambia-HPTN 071 (PopART). Health Policy Plan [Internet]. 2021;36(6):881–90. Available from: https://researchonline.lshtm.ac.uk/id/eprint/4660823/1/Lario-healthworkers-PopART.pdf

23. Shanaube K, Schaap A, Floyd S, Phiri M, Griffith S, Chaila J, et al. What works –reaching universal HIV testing: lessons from HPTN 071 (PopART) trial in Zambia. AIDS [Internet]. 2017 Jul 17;31(11):1555–64. Available from: http://www.ncbi.nlm.nih.gov/pmc/articles/PMC5491236/

24. Bond V, Hoddinott G, Viljoen L, Ngwenya F, Simuyaba M, Chiti B, et al. How ‘place’ matters for addressing the HIV epidemic: evidence from the HPTN 071 (PopART) cluster-randomised controlled trial in Zambia and South Africa. Trials [Internet]. 2021;22(1):1–13. Available from: https://trialsjournal.biomedcentral.com/track/pdf/10.1186/s13063-021-05198-5.pdf

25. LaMorte WW. Screening for Disease [Internet]. Boston University School of Public Health. 2016 [cited 2018 Oct 4]. Available from: http://sphweb.bumc.bu.edu/otlt/MPH-Modules/EP/EP713_Screening/EP713_Screening-TOC.html

26. WHO. HIV and Adolescents: Guidance for HIV Testing and Counselling and Care for Adolescents Living with HIV: Recommendations for a Public Health Approach and Considerations for Policy-Makers and Managers [Internet]. Geneva; 2013. Available from: https://www.ncbi.nlm.nih.gov/books/NBK217962/pdf/Bookshelf_NBK217962.pdf

27. World Health Organization. HIV and Adolescents: Guidance for HIV Testing and Counselling and Care for Adolescents Living with HIV. Recommendations for a Public Health Approach and Considerations for Policy-Makers and Managers. WHO Publ. 2013;100.

